# The striatum is an early, accurate indicator of amyloid burden using [^11^C]PiB in Down syndrome: comparison of two radiotracers

**DOI:** 10.1101/2024.12.04.24318526

**Authors:** Max McLachlan, Brecca Bettcher, Andrew McVea, Alexandra DiFillipo, Matthew Zammit, Lisette LeMerise, Jeremy Rouanet, Julie Price, Dana Tudorascu, Charles Laymon, David Keator, Patrick Lao, Adam M. Brickman, Tim Fryer, Sigan Hartley, Beau M. Ances, Sterling Johnson, Tobey Betthauser, Charles K. Stone, Shahid Zaman, Benjamin Handen, Elizabeth Head, Mark Mapstone, Bradley T. Christian

## Abstract

**INTRODUCTION:** Adults with Down syndrome demonstrate striatum-first amyloid accumulation with [^11^C]PiB PET imaging, which has not been replicated with [^18^F]florbetapir (FBP). Early striatal accumulation has not been temporally quantified with respect to global cortical measures.

**METHODS:** Longitudinal PiB (n=175 participants) and FBP (n=92 participants) data from the Alzheimer Biomarkers Consortium-Down Syndrome were used to measure cortical and striatal binding. Generalized temporal models for cortical and striatal amyloid accumulation were created using the sampled iterative local approximation (SILA) method.

**RESULTS:** PiB demonstrated greater striatal-to-cortical ratios than FBP. SILA analysis revealed striatal amyloid burden occurs 3.40 (2.39) years earlier than the cortex in PiB. There was no difference between the cortex and striatum in FBP.

**DISCUSSION:** Among adults with Down syndrome, the striatum consistently accumulates amyloid earlier than the cortex when measured with PiB. This suggests the striatum is more sensitive to the onset of PiB PET-detectable amyloid in Down syndrome.

## 1. Introduction

Down syndrome (DS) is a developmental disability caused by the triplication of chromosome 21. People with DS carry a 90% lifetime risk of developing Alzheimer’s Disease (AD)^1^, caused by the overproduction of amyloid precursor protein expressed on chromosome 21^2^. Nearly all adults with DS over the age of 40 years exhibit protein neuropathologies associated with AD, including amyloid plaques and neurofibrillary tau tangles^3^, and the median age for developing clinical symptoms is estimated to be between 50-55 years^4^. Consequently, AD is one of the leading causes of death for adults with DS and characterization of neuropathological features is necessary to improve long-term health outcomes in this at-risk population^5^.

Amyloid-targeting immunotherapies are emerging treatments for mitigating amyloid plaque accumulation and reducing cognitive impairment associated with late onset AD^6^. Early clinical trials for these therapies excluded individuals with lifelong cognitive dysfunction, including those with DS, and limited the age criteria to older cohorts (> 50 or 60 years)^7–9^. Additionally, the excess production and accumulation of amyloid in the DS population is associated with significant cerebral amyloid angiopathy (CAA), which can cause complications such as amyloid-related imaging abnormalities (associated with inflammation or stroke)^10^. These restrictions prevented the DS population from participating in anti-amyloid drug trials^11–13^. To address these concerns, multiple trial-ready cohorts have recently been established for the DS population^14,15^, and criteria changes have been proposed to the FDA and drug formularies^16,17^. To provide the same clinical trial opportunities to people with DS and mitigate potential adverse responses, early amyloid detection and accurate disease staging is of critical importance^18^.

The earliest amyloid (Aβ) detection in the DS population is in the striatum based upon studies using PET radiotracer [^11^C]PiB. This striatum-first pattern was first observed in autosomal-dominant forms of AD (ADAD)^19^ – these studies found both greater overall tracer binding in the striatum and earlier binding than the global cortex relative to disease stage^19–23^. This pattern was generally universal across ADAD mutation types, though heterogeneity is observed^23^. Striatum-first accumulation in DS was previously reported in longitudinal studies^24–26^, with detectable accumulation at approximately 40 years of age. These findings are corroborated when DS and ADAD cohorts are compared with sporadic AD^27,28^. When compared longitudinally with the global cortex, the striatum shows significantly earlier accumulation but no difference in the rate of accumulation^29^. Thus, the striatum may serve as an earlier indicator of amyloid burden in the DS population than conventional global measures, such as Centiloids^30^. However, the extent and magnitude of early striatal amyloid accumulation has not been directly quantified relative to the cortex, limiting its current feasibility for research and clinical use.

The striatum-first pattern of amyloid deposition is exclusively observed using the radiotracer [^11^C]PiB in both the DS^24–29,31,32^ and ADAD^19–23,33^ populations. Although striatal binding is consistently reported alongside cortical binding using [^18^F]florbetapir (FBP) in cross-sectional studies of ADAD and DS^34–37^, striatum-first accumulation has yet to be observed longitudinally or between cognitive groups with this radiotracer^37–40^. For early striatal amyloid accumulation to be a clinically useful measure of AD staging in DS, its prevalence must be confirmed across PET radiotracers used in the DS population.

This study examined longitudinal changes to amyloid PET signal in the striatum using PiB and FBP in DS cohorts. All participants were recruited through the Alzheimer Biomarkers Consortium – Down Syndrome (ABC-DS), a large multi-site study investigating AD-related pathology in adults with DS^14^. To produce general models for the longitudinal change of Aβ PET in these regions, the sampled iterative local approximation (SILA) algorithm was implemented. SILA is used to combine large longitudinal amyloid PET datasets in both the late onset AD and DS populations^41,42^. SILA’s estimates for age-of-pathology-onset are particularly useful for staging AD pathologies temporally. In this study, the SILA method was applied to both cortical and striatal PET data, allowing for the direct comparison of the estimated age-of-onset in each region.

## 2. Methods

### Participants

At four ABC-DS sites, 175 participants (47.4% female; average age at baseline visit = 37.1 (8.1) years) underwent up to six PET PiB imaging scans. At three ABC-DS sites, a separate group of 97 participants (35.9% female; average age at baseline visit = 50.1 (7.2) years) underwent up to four PET FBP imaging scans. Time between longitudinal PiB scans was approximately 3.2 (0.9) years, while time between longitudinal FBP scans was approximately 1.6 (0.8) years. All participants received structural T1-weighted imaging at each timepoint. Blood samples were collected at each timepoint to determine APOE allele status. The study was approved by an Institutional Review Board and conducted in accordance with the Declaration of Helsinki. Informed consent and assent were obtained prior to conducting any study activities. The enrollment criteria for PiB sites included being ≥ 25 years of age and having a non-verbal mental age of at least 3 years, based on the Stanford-Binet, 5th edition. The enrollment criteria for the FBP sites included being ≥ 40 years of age, resulting in an older cohort relative to the PiB cohort. All participants received genetic testing to confirm DS status by karyotype. Exclusion criteria restricted those having an unstable psychiatric or medical condition that impaired cognition or contraindicated the acquisition of brain imaging scans. See Table 1 for information about longitudinal scanning and participant demographics.

**Table 1.** Participant demographics for PiB (top) and FBP (bottom) cohorts. Amyloid positivity was determined using a cutoff of 18.1 Centiloids (CL). Aβ converters have at least one Aβ− scan followed by an Aβ+ scan. No participants revert from Aβ+ to Aβ−.

| | 1 Scan | 2 Scans | 3 Scans | $\geq 4$ Scans | All Participants |
| --- | --- | --- | --- | --- | --- |
| **PiB** |  |  |  |  |  |
| **Cohort Size (n)** | 64 | 59 | 15 | 37 | 175 |
| **Age at Baseline, mean (SD) (yrs)** | 39.5<br>(SD = 9.1) | 35.0<br>(SD = 7.4) | 37.4<br>(SD = 6.9) | 36.0<br>(SD = 6.4) | 37.1<br>(SD = 8.0) |
| **Female (%)** | 28 (43.8%) | 32 (54.2%) | 6 (40.0%) | 17 (46.0%) | 83 (47.4%) |
| **Aβ- (n)** | 46 | 45 | 6 | 16 | 113 |
| **Aβ Converter (n)** | N/A | 7 | 5 | 17 | 29 |
| **Aβ+ (n)** | 18 | 7 | 4 | 4 | 33 |
| **FBP** |  |  |  |  |  |
| **Cohort Size (n)** | 25 | 29 | 29 | 9 | 92 |
| **Age at Baseline, mean (SD) (yrs)** | 52.8<br>(SD = 7.6) | 49.1<br>(SD = 7.4) | 49.3<br>(SD = 6.6) | 48.7<br>(SD = 6.7) | 50.1<br>(SD = 7.2) |
| **Female(%)** | 11 (44.0%) | 12 (41.4%) | 7 (24.1%) | 3 (33.3%) | 33 (35.9%) |
| **Aβ- (n)** | 7 | 4 | 4 | 1 | 16 |
| **Aβ Converter (n)** | N/A | 0 | 6 | 2 | 8 |
| **Aβ+ (n)** | 18 | 25 | 19 | 6 | 68 |

**Table 2.** Parameter estimates and significance for multivariate linear model. For Cohen’s f: f = 0.10 is the suggested cutoff for a small effect size (S) f = 0.25 is the suggested cutoff for a medium effect size (M) f = 0.40 is the suggested cutoff for a large effect size (L)

| Model Term | $\beta$<br>[95% C.I.] | P | Cohen's $f$ |
| --- | --- | --- | --- |
| Intercept | 0.711<br>[0.534, 0.887] | N/A | N/A |
| CL | 5.206e-3<br>[3.094e-3, 7.317e-3] | <0.001*** | 0.44 (L) |
| CL<sup>2</sup> | -4.426e-5<br>[-6.226e-5, -2.625e-5] | <0.001*** | 0.44 (L) |
| Tracer | **0.210**<br>**[0.155, 0.264]** | **<0.001\*\*\*** | **0.64 (L)** |
| APOE4 | 0.174<br>[-0.179, 0.527] | 0.018* | 0.22 (S) |
| Sex | 3.946e-2<br>[-8.040e-3, 8.695e-2] | 0.102 | 0.15 (S) |
| Age | 3.994e-3<br>[1.385e-3, 7.850e-3] | 0.073 | 0.16 (S) |
| **APOE4:Tracer** | -0.163<br>[-0.276, -0.049] | 0.006** | 0.25 (M) |
| **APOE4:Age** | -3.505e-3<br>[-1.064e-2, 3.634e-3] | 0.333 | 0.09 |

### MRI Imaging

T1-weighted magnetic resonance images (MRI) were acquired on the following scanners, depending on the imaging site: 3T GE Discovery MR750, Siemens MAGNETOM Prisma^Fit^, GE Signa PET/MR, Siemens Biograph mMR, and Phillips Achieva. T1-weighted scans were acquired with one of the following sequences: 3D Fast Spoiled Gradient Echo (FSPGR) or Magnetization-Prepared Rapid Gradient-Echo (MP-RAGE). Images were corrected for magnetic field bias and gradient non-linearity, and a quality control procedure evaluated images for any remaining artifacts.

### PET Imaging

PiB PET scans (target injected dose of 15 mCi) were acquired on the following scanners, depending on the imaging site: ECAT EXACT HR+, SIGNA PET/MR, Biograph 64 mCT, Discovery 710, Biograph 40 True Point. FBP PET scans (target injected dose of 10 mCi) were acquired on the following scanners, depending on the imaging site: HRRT, Biograph 64 mCT, Biograph mMR, Biograph 64 mCT 4R. PET data was collected 50-70 minutes post-intravenous injection for both PiB and FBP, resulting in four 5-minute frames for each scan.

### Image Processing

All image post-processing was performed using statistical parametric mapping software (SPM12). Individual T1-weighted MRIs underwent tissue classification using SPM12 Segmentation. The brain was skull-stripped by combining the white matter (WM), grey matter (GM), and cerebrospinal fluid (CSF) tissue segmentations as follows: WM + GM + 0.25*CSF (CSF is included to improve normalization to ventricles and other low intensity regions in the MRI). An eroded white matter (EWM) mask was produced from each MRI by smoothing the WM segmentation to 8 mm resolution and thresholding to 90% tissue probability. This produced participant-specific masks containing the centrum semiovale, pons, cerebellar white matter, and corpus callosum.

PET images were smoothed to 8 mm isotropic resolution, accounting for PET scanner resolution and site-specific smoothing. Images were summed 50-70 minutes and co-registered to the participant’s T1-weighted MRI using SPM12 Coregister. The skull-stripped MRI was normalized to a DS-specific template MRI (LeMerise et al; AAPM 2022) using the SPM12 Old Normalize function. The same transformation matrix was applied to the co-registered PET scan to warp them into a common imaging space.

Global amyloid burden was calculated following the Centiloid (CL) calibration procedure^30^. The CL global cortical ROI, whole cerebellum (WC) ROI, and the Harvard Oxford Atlas were warped into the DS template space using SPM12 Old Normalize. The warped ROIs were smoothed by 2mm to improve margin uniformity. For PiB, SUVR^50-70^ parametric images were created using the WC as reference region^43^. For FBP, the WC and white matter regions are used as reference regions in the NT population – most studies report the greatest power and longitudinal stability when using white matter-based reference regions^44,45^. To determine the optimal reference region for longitudinal comparison in the DS population, SUVR^50-70^ parametric images were created for each FBP scan using both the WC and participant-specific EWM as reference regions. Stability of longitudinal trajectories determined which reference region was used for the remaining analyses in all FBP scans. A CL calibration was then performed for both the PiB and FBP processing methods^30^.

Average cortical SUVR was converted to CL. Participants were initially grouped by amyloid positivity status using the published threshold of 18.1 CL^24^. An average PET image was created for each group. A PET-generated striatum ROI was then created by subtracting the average Aβ− PiB image from the average Aβ+ PiB image, thresholding to a difference of +0.75 SUVR units, and removing unassociated regions. SUVR was calculated for the caudate, putamen, accumbens, composite MRI-based striatum (all made using the Harvard Oxford Atlas), and the PET-generated striatum.

### Statistics

For the FBP reference region analysis, the average rate of cortical SUVR change was assessed with % change/year = (SUVR_2_-SUVR_1_)/(SUVR_1_*Δt). The number of scans with positive longitudinal changes was counted for each reference region. SUVR was assessed using a linear mixed effects (LME) model of the form SUVR ~ Scanner (4 different scanners) + Time (relative to first scan) + Reference Region, with a random participant-level intercept. A paired t-test and Levene’s test evaluated differences in the distributions of % Change/year for each reference region method. Group differences in white matter-to-cerebellum ratios are also evaluated between PiB and FBP groups.

For each PET scan, the ratio of striatum SUVR-to-cortex SUVR was calculated. To minimize group-level differences in Aβ status, scans were Centiloid matched within +/− 5 CL across tracers. Only one scan per participant was permitted to be matched. The data from these participants were incorporated into a linear model of the form Ratio ~ CL + CL^2^ + Tracer + APOE4 + Age + Sex + APOE4:Tracer + APOE4:Age. Significance of model terms was evaluated using Type 2 ANOVA. Effect size was calculated using Cohen’s f statistic for each model term.

Positivity thresholds were calculated for all striatum-specific ROIs and the cortical ROI (for consistency) using a gaussian mixture model with 2 distributions. The thresholds were defined as 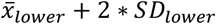^46^. Following the SILA method, the average rate of change was calculated within discrete SUVR intervals. Defining t = 0 as the time of positivity, Euler’s method determines the average longitudinal change. This process yields generalized models for the progression of SUVR relative to positivity in each ROI. The estimated time-to-amyloid-positivity for each participant and ROI was calculated by minimizing the sum of squared differences between participant trajectories and the models. Paired t-tests evaluated the average differences in time-to-positivity between the cortex and each striatum-specific ROI. Estimates from SILA are most reliable for trajectories that have crossed the positivity threshold; therefore, this analysis was only performed for participants with at least one positive scan in each ROI.

Each SILA model is plotted relative to the individual ROI’s time-to-positivity axis, meaning the models do not inherently exist on the same time axis. In order to compare the striatum model to the cortex model, the cortex model is shifted by the average time difference found from the paired t-tests.

Following this transformation, the striatum-to-cortex model ratio can be calculated.

## 3. Results

### FBP Reference Region Comparison

Figure 1 displays FBP cortical longitudinal trajectories and % change/year between timepoints using WC and EWM reference regions. With the WC reference region, 67/114 (59%) consecutive scans show longitudinal increases in amyloid, with the average rate of change being 1.92 ± 6.04 %/year.

**Figure 1.**
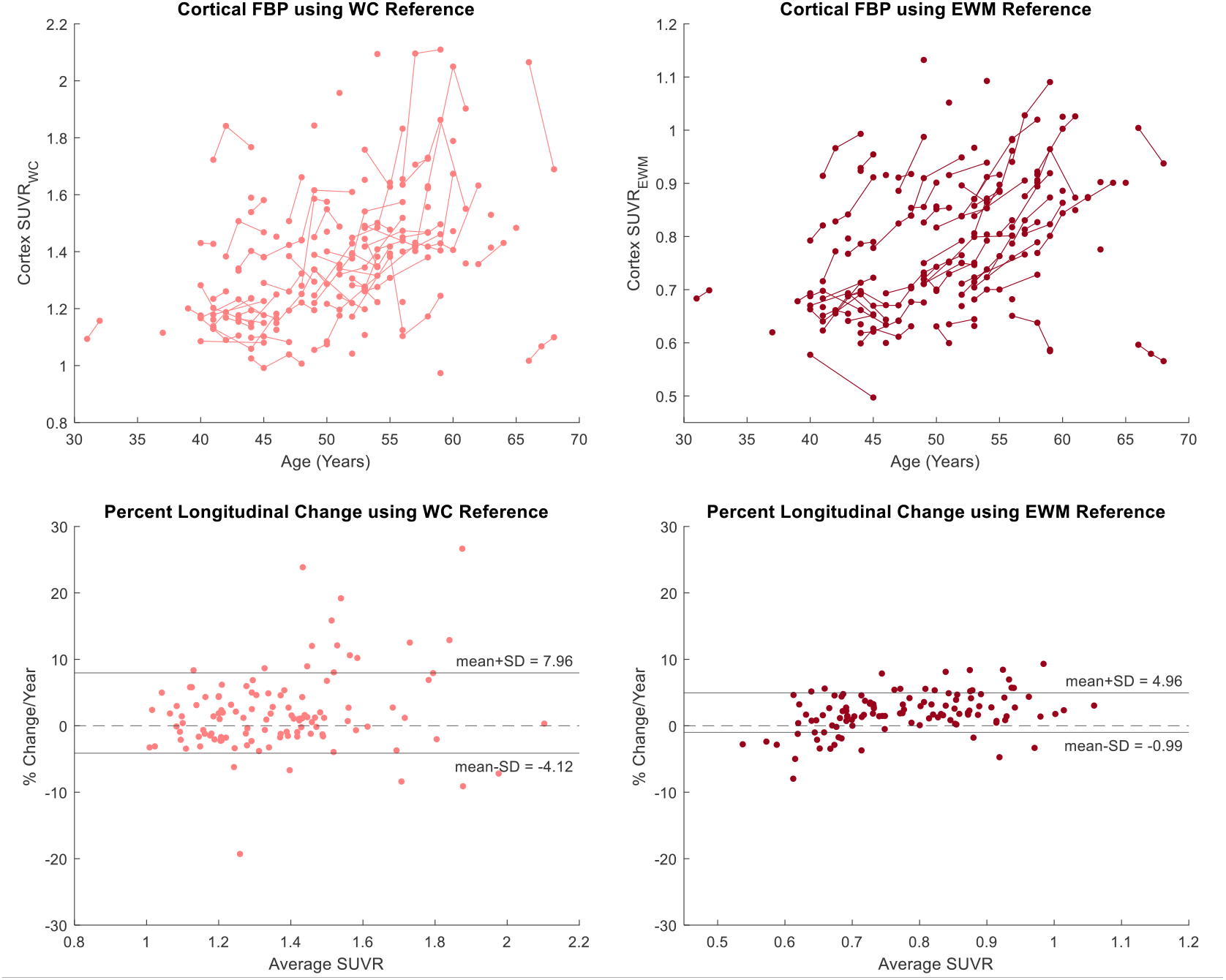
Longitudinal FBP trajectories (top) and percent longitudinal change (bottom) using EWM and WC reference regions showing that using EWM provides more sensitivity and less variability. SUVREWM has a lower and compressed range when compared to SUVRWC. Percent longitudinal change was calculated as follows: % change/year = (SUVR_2_-SUVR_1_)/(SUVR_1_*Δt).

With the EWM reference region, 94/114 (82%) consecutive scans show longitudinal increases, with the average rate of change being 1.98 ± 2.97 %/year. Based on the LME model, SUVR_EWM_ is lower than SUVR_WC_ (p<0.001***). All model estimates are provided in Supplementary Material. A paired t-test for % change/year showed no significant difference in means between reference region methods (p=0.909). Levene’s test showed a significant reduction in variance when using the EWM reference region, with an estimated variance ratio σ^2^_EWM_/σ^2^_WC_ = 0.259 [0.167,0.351] (p<0.001***). As the EWM reference region method showed a greater number of participants with longitudinal increases and lower group variance, SUVR_EWM_ was used for the remaining analysis with all FBP scans. A CL calibration is included in Supplementary Material for both PiB and FBP.

### PiB/FBP Comparison

Figure 2 displays longitudinal trajectories for PiB and FBP in both the cortex and PET-generated striatum ROIs. Figure 3 shows average Aβ− and Aβ+ images for each radiotracer, as well as % difference images between amyloid groups. Both tracers demonstrated the highest change in amyloid in cortical regions, particularly the precuneus and striatum. PiB showed greater overall range of % difference, likely influenced by the greater dynamic range of SUVR_WC_ compared to SUVR_EWM_.

**Figure 2.**
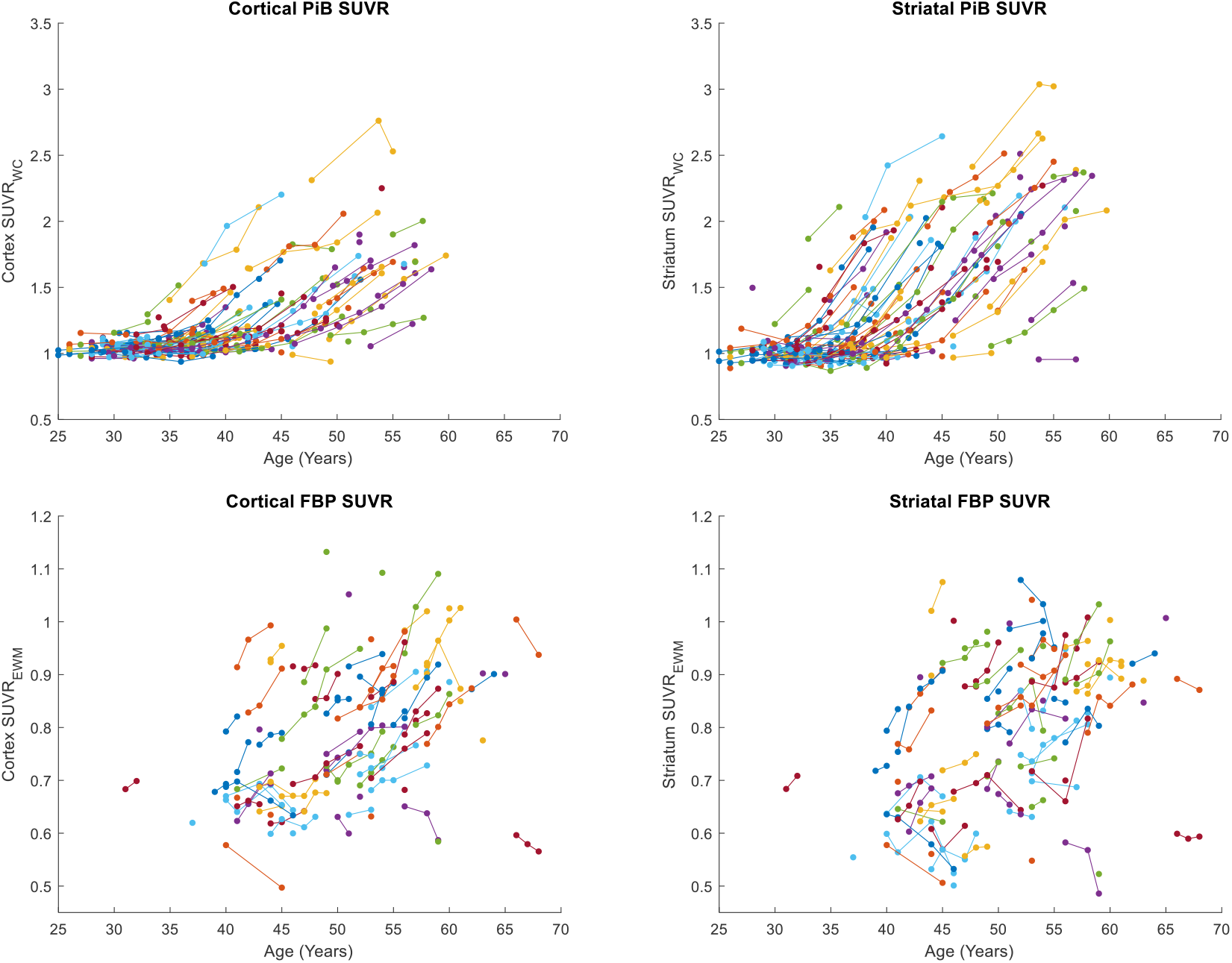
Longitudinal PiB (top) and FBP (bottom) trajectories for both the global cortex and PET-generated striatum ROI. Individual participants are denoted by marker color.

**Figure 3.**
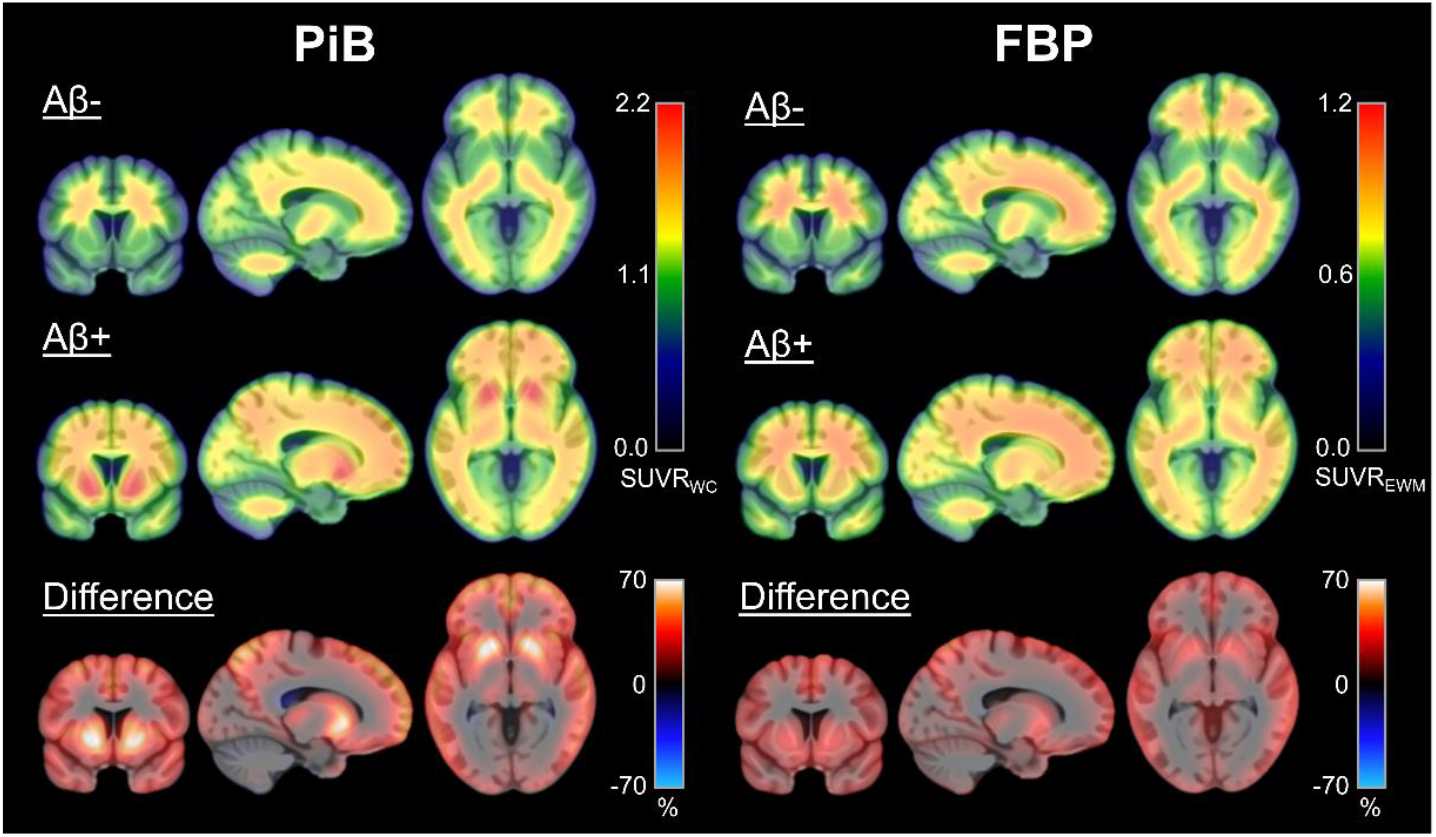
Average PET SUVR images by amyloid status. Percent difference image shows the greatest PET signal in the precuneus and striatum for both tracers, though PiB has a greater dynamic range and shows much higher relative signal than FBP.

CL matching across tracers resulted in 66 matched pairs (demographics in Suplementary Materials). The groups showed significant sex and age differences. These scans were used for the linear model Ratio ~ CL + CL^2^ + Tracer + APOE4 + Age + Sex + Age:Sex + APOE4:Tracer + APOE4:Age (Figure 4). Participants who were imaged with PiB show a significantly higher striatum-to-cortex ratio than those imaged with FBP (p<0.001***), with the modeled ratio maximizing at approximately 75 CL. The Tracer model term shows the largest effect size, followed by CL terms and APOE4 status (Table 4).

**Figure 4.**
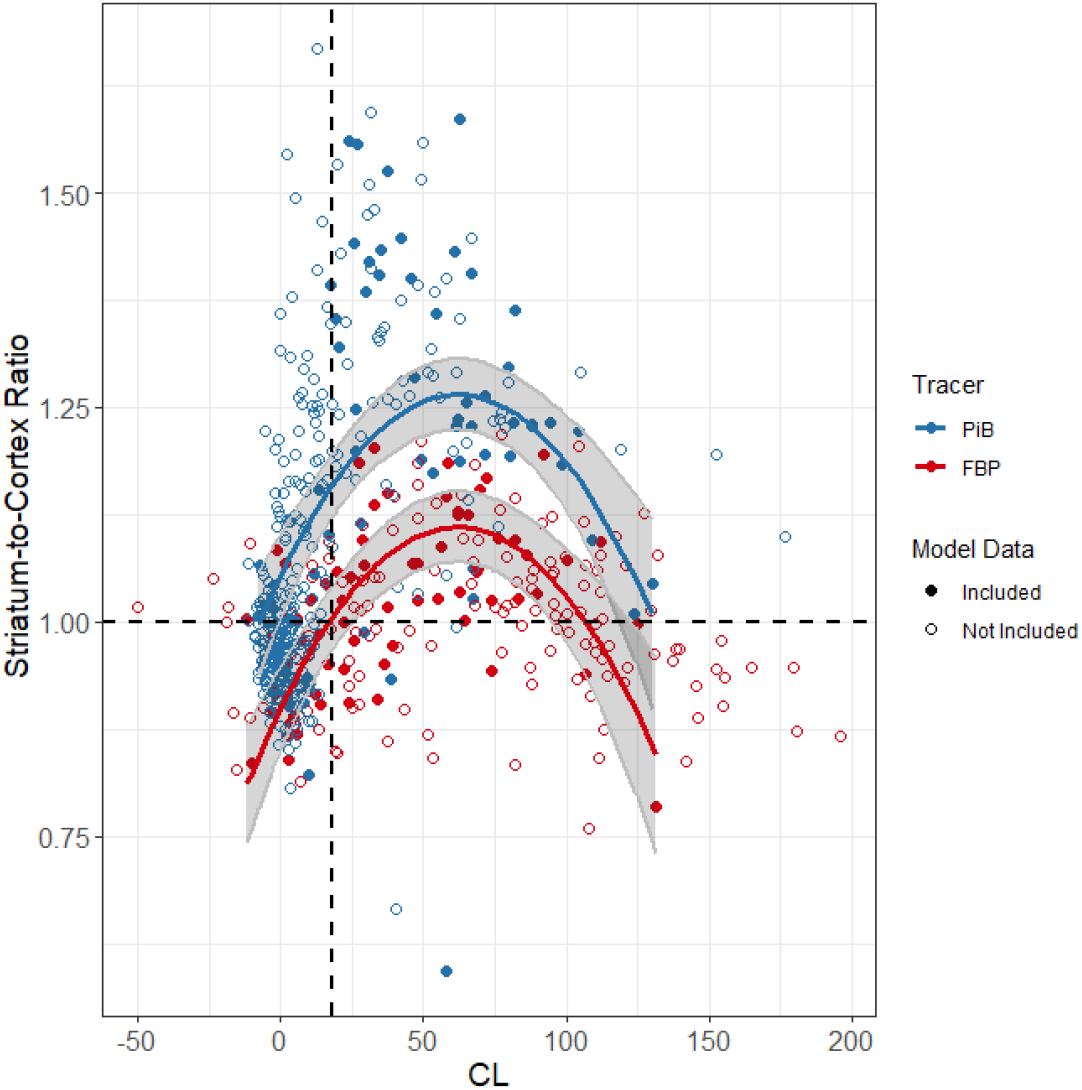
Multivariate linear model for the striatum-to-cortex ratio. Centiloid (CL)-matched scans (included in the model) are denoted by solid markers. For visual simplicity, the displayed model is of the form Ratio ~ CL + CL^2^ + Tracer.

Across the entire cohort, PiB had higher SUVR in the striatum than the cortex, with an average difference of 0.13 SUVR (two-sample t-test, Cohen’s d = 0.35, p<0.001***). FBP showed an average difference between striatum SUVR and cortex SUVR of 0.01 (two-sample t-test, Cohen’s d = 0.04, p=0.155). Additionally, FBP showed a singificantly higher white matter-to-cerebellum ratio on average (1.76 (0.14)) compared to PiB (1.63 (0.11)) across the entirety of both cohorts (two-sample t-test, Cohen’s d = 1.05; p<0.001***).

### SILA Analysis

SILA modeling was completed on the cortex and all striatum-specific regions using GMM-derived cutoffs (listed in Supplementary Materials). In general, all Aβ-converting participants have a striatum positive timepoint before their first cortex positive timepoint in the PiB cohort. For each participant with at least one positive scan, the estimated time-to-cortex-positivity was subtracted from the estimated time-to-positivity in each striatum-specific region (Figure 5). For PiB, the PET-based striatum showed the earliest positivity estimates, on average 3.40 (2.39) years before the cortex. The caudate showed the latest positivity estimates, on average 4.22 (10.98) years after the cortex. Other than the caudate, all striatum ROIs showed earlier time-to-positivity estimates than the cortex in the PiB cohort. For FBP, the PET-based striatum, putamen, and accumbens ROIs did not show different time-to-positivity estimates than the cortex. SILA models could not be generated for the caudate and MRI-based striatum ROIs, as high longitudinal variability in these regions resulted in a non-monotonic model.

**Figure 5.**
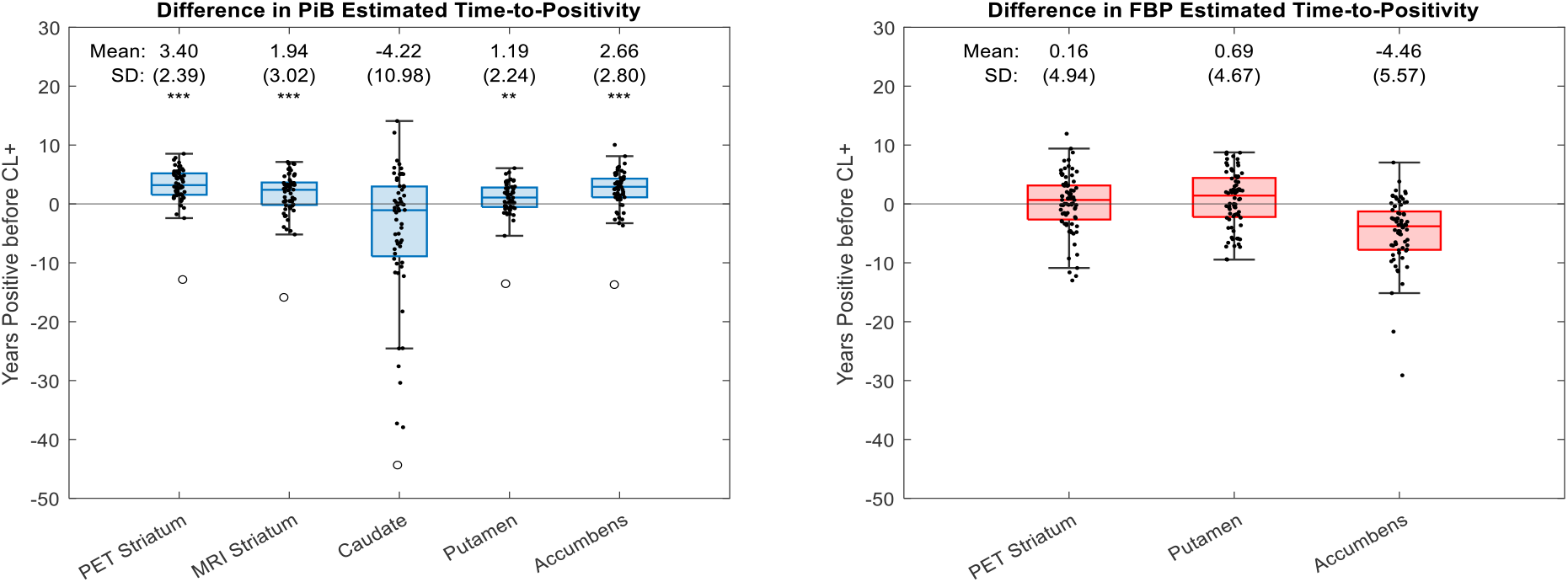
Difference in estimated time-to-positivity between the cortex and each striatum-specific ROI. ROIs include the caudate, putamen, accumbens, and a composite MRI-based striatum, all generated using the Harvard Oxford Atlas. Additionally, a PET-generated striatum ROI was created by thresholding the difference image in Figure 1a to +0.75 SUVR. Differences in estimated time-to-positivity relative to the cortex were assessed with a one-tailed paired t-test, determining if each region shows earlier estimates than the cortex. PiB (left) shows significantly earlier amyloid positivity in all regions except the caudate. FBP (right) does not show any significant difference with the cortex. A monotonically increasing SILA model could not be created for the caudate and MRI-based striatum ROIs in FBP due to longitudinal variability. One individual in the PiB cohort was deemed an outlier with a Z score < −4 in all ROIs.

To determine the striatal-to-cortical ratio relative to time-to-positivity in PiB, the cortical SILA curve was shifted by the average calculated difference (3.40 years), aligning the model on the PET-based striatum positivity axis (Figure 6). At the time of striatum positivity, the ratio of the two SILA models was 1.03 [1.01, 1.06]. At the time of cortical positivity, the ratio of the two SILA models was 1.16 [1.13,1.20].

**Figure 6.**
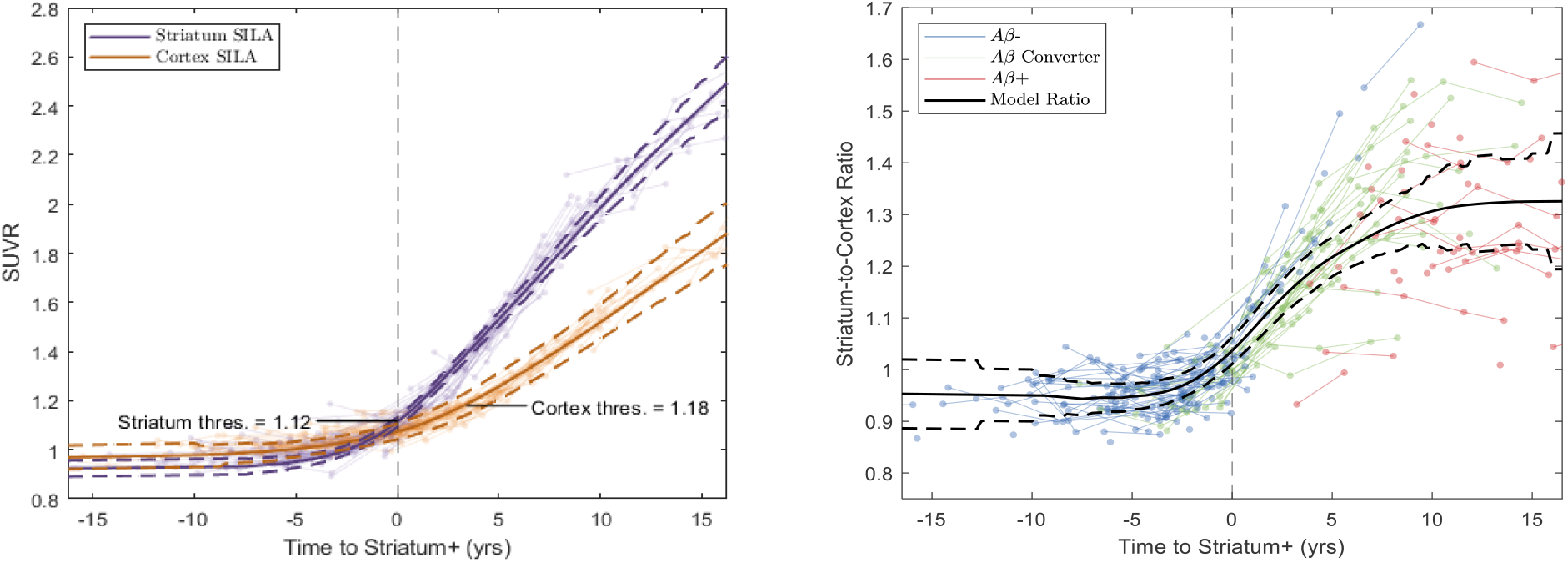
SILA models for the cortex and PET-based striatum ROIs of PiB (left). To plot the cortex model along the striatum positivity axis, the cortex model was shifted by the average difference found in Figure 5 (3.40 years). Confidence bands were adjusted accordingly. Participant trajectories for each ROI are displayed as semi-transparent markers. Ratio of the striatum SILA model to the cortex SILA model is displayed right. Participant trajectories are color-coded by cortical amyloid status. All cortically Aβ− converting participants demonstrate earlier striatal positivity.

## 4. Discussion

With the development and implementation of anti-amyloid therapies, early detection of AD pathology is critical for identifying appropriate inclusion of people with DS in clinical trials^18^. This study supports previous findings that the striatum is the earliest region of amyloid accumulation detectable using [^11^C]PiB in people with DS. Furthermore, FBP is insensitive to striatum-first accumulation in DS, suggesting there exists a fundamental difference in the binding characteristics between PiB and FBP in the striatum.

The exact mechanism through which PiB detects early striatal accumulation is unknown. Post-mortem histopathological studies demonstrated the presence of diffuse Aβ plaque in the striatum for both ADAD^47–49^ and DS^50–52^. These diffuse plaques are largely formed from Aβ42 peptides rather than Aβ40^48,53^. High binding affinity for Aβ42 neuritic plaques is well documented for both PiB^54,55^ and FBP^56,57^. Binding to diffuse plaque is less understood, which likely contributes to discrepancies observed between post-mortem pathology and in vivo PET measures^58^. Neuropathology studies show amyloid first appears in the frontal cortex up to 20 years before the striatum^59,60^, with a period of rapid amyloid accumulation around 30-40 years of age in DS^60^. Of note, soluble protofibrillar and oligomeric forms of amyloid are elevated before the formation of plaques throughout the brain in DS^61–64^. Similar distributions were observed in various ADAD mutations^62^. These studies are not specific to the striatum, but the rapid elevated presence of diffuse plaque in the striatum is logically preceded by smaller subunits^65^. PiB binds to oligomeric and larger protofibrillar forms of amyloid, in addition to diffuse plaque (although this binding is weaker when compared to neuritic fibrillar forms)^47,54,55^. Significant FBP binding to high molecular weight protofibrils but weaker binding to smaller protofibrils and oligomers is observed^66,67^. Post-mortem studies with FBP do not comment on binding differences between diffuse plaque and neuritic plaque scores^35,56,57,68^. These differences between PiB and FBP binding to amyloid subtypes may explain the observed discrepancy in the striatum-first amyloid binding in DS. A direct in vitro comparison of the binding affinities of PiB and FBP to diffuse plaque could clarify this difference.

Eroded WM reference region normalization (i.e. uptake ratio) demonstrates greater longitudinal stability than the WC with FBP. These results are consistent with previous findings that normalization to a high intensity white matter reference region increases the power of longitudinal estimates with FBP^44,45^. Additionally, FBP shows significantly higher white matter-to-cerebellum ratios than PiB. PET studies in animal models showed that FBP may bind more strongly to myelin than PiB^69^. However in vivo studies suggest that lower relative cortical binding and a smaller dynamic range in FBP may explain the apparent elevated white matter binding^70,71^. Therefore, partial volume effects from white matter retention may influence the variability of longitudinal measurements in FBP^72^. As the main outcome measure of the current study was the ratio of striatal to cortical signal, partial volume effects are likely minimal.

According to SILA analysis in the PiB cohort, the caudate revealed lower sensitivity to amyloid accumulation and greater variability in estimated time-to-onset than other striatal subregions. Pathological estimates suggest that amyloid plaque develops ubiquitously throughout both the caudate and putamen^53^, so this observation is likely influenced by spatial smoothing or imperfect normalization of atrophied ventricles^73^.

The striatum-specific region that showed the earliest amyloid accumulation was generated by thresholding the PiB difference image between Aβ groups. This region comprised the accumbens, putamen, and inferior ventral caudate. Longitudinal SUVR in this region demonstrated nearly universal early positivity and higher signal in Aβ-converting participants relative to Centiloids. A significant interaction between APOE status and tracer is observed, which may be influenced by the difference in age range between the two tracer cohorts. One individual was deemed an outlier in the SILA analysis, as they demonstrated a Z score < −4 in all ROIs investigated and was amyloid positive in the cortex but not in the striatum for all scans. Interestingly, this participant possesses a mosaic karyotype of DS, meaning only a percentage of their cells exhibit trisomy 21. The impact of mosaicism for trisomy 21 is an active area of study.

There are two main limitations to this work. First, the use of a common template space for image spatial normalization may impact PET estimates for participants with severe structural atrophy (roughly 10% of participants). As mentioned, the striatum-to-cortex outcome measure is likely more robust to atrophy and partial volume effects than SUVR measures alone. Future analyses can be performed using native space regional parcellations to reduce these potential effects. Second, the same longitudinal data used in the creation of the SILA models was also used for the estimates of time-to-positivity. This approach may bias the time-to-positivity estimates in areas of a model with few constituent trajectories. Differences in age and scanning interval between tracer cohorts may additionally bias the results. As the paired t-test used within-participant comparisons, the significance of the time-to-positivity estimates is likely unaffected by these confounds.

Future work should investigate the striatum-first pattern using alternative amyloid radiotracers. The speculated binding differences between FBP and PiB may be relevant for all stilbene-based amyloid tracers. One study using stilbene tracer [^18^F]florbetaben reported moderately elevated striatal uptake in an ADAD cohort^74^. A case study of an individual with early-onset AD also found high florbetaben binding in the striatum^75^. However, these studies were not longitudinal and did not demonstrate the same universal pattern seen with PiB. In particular, a comparison of PiB with other benzothiazole derivatives (e.g. [^18^F]flutemetamol, [^18^F]NAV-4694) may clarify the differential sensitivity of this family of radiotracers to striatal accumulation. With the availability of a cyclotron limiting many clinical trial screening sites to F-18 radiotracers, the tracer dependence of striatum-first amyloid accumulation may determine its feasibility as a useful clinical marker.

When comparing amyloid PET tracers PiB and florbetapir, only PiB detects early striatal accumulation in DS, with striatal positivity occurring on average 3.40 years earlier than cortical positivity. These results suggest that the striatum should be used alongside cortical measures as an earlier indicator of amyloid pathology in DS. Understanding the differences in amyloid binding between radiotracers is important for directing upcoming clinical AD trials in DS, for which the assessment of early amyloid burden is a primary treatment target.

## Data Availability

All data produced in the present work are contained in the manuscript.

## 5. Acknowledgements

This work was supported by NIH funding U01 AG051406, U19 AG068054, P50 HD105353, P30 AG062715, P50 AG005133, RF1 AG025516 and R01 AG031110. Special thanks to the participants and their families who took part in these studies. The authors have no conflicts of interest to disclose.

