## SupplementaryMaterial for "The striatum is an early, accurate indicator of amyloid burden using [^11^C]PiB in Down syndrome: comparison of two radiotracers"

**Supplementary Material**

Linear Mixed Effects Model for FBP Reference Regions

| Model Term | β<br>[95% C.I.] | P | Cohen's f |
|---|---|---|---|
| **Intercept** | 1.357<br>[1.306, 1.408] | N/A | N/A |
| **Time** | 0.017<br>[0.010, 0.024] | <0.001*** | 0.27 (M) |
| **Reference Region** | -0.589<br>[-0.606, -0.572] | <0.001*** | 3.88 (L) |
| **Scanner** | | | |
| Scanner 1 | 0.0301<br>[-0.046, 0.106] | 0.792 | 0.06 |
| Scanner 2 | -0.045<br>[-0.233, 0.143] | | |
| Scanner 3 | -0.065<br>[-0.127, -0.003] | | |

Table S1 – LME for SUVR calculated using EWM and WC reference regions.

<u>Centiloid Calibration</u>
Level 1: Replication Analysis

All Level 1 calibration results are within tolerance (see Table S1), so a Level 2 calibration can be performed.

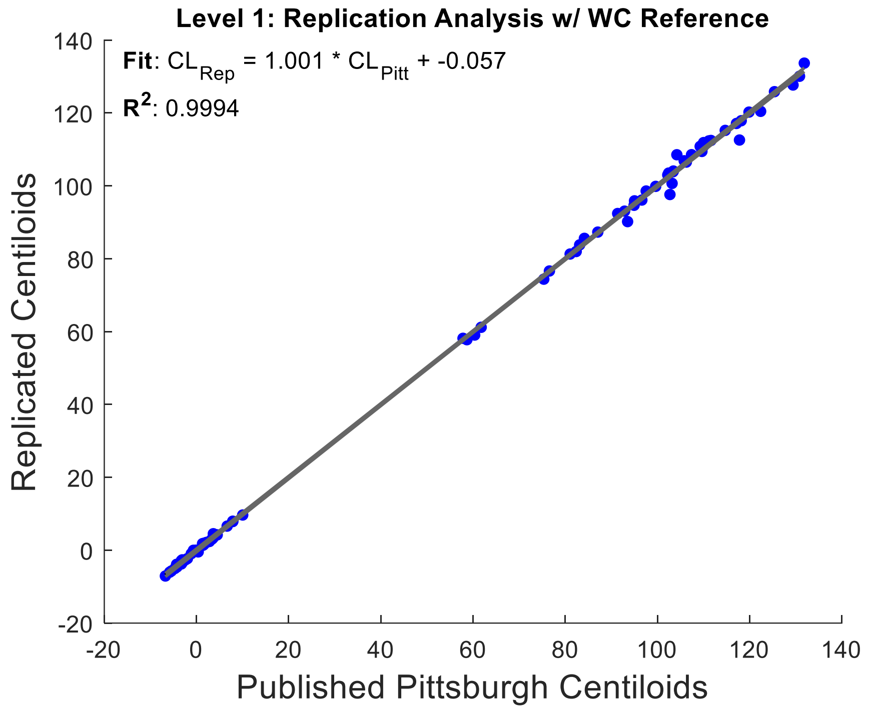


Figure S1 – Level 1 Centiloid replication analysis using published Pittsburgh calibration data.

| Reference VOI | Slope (0.98 to 1.02) | Intercept (-2 to 2 CL) | $R^2$ (> 0.98) | % Difference YC (-2 to 2%) with SD ± 2% |
|---|---|---|---|---|
| **Whole Cerebellum** | 1.001 | -0.057 CL | 0.999 | -0.3 ± 0.4 % |

Table S2 – Regression information for Level 1 Centiloid replication analysis. All results are within tolerance as listed in column titles.

For PiB, the calibration used the published Pittsburgh data included with the Centiloid Project. For FBP, the calibration used published Avid data, which included 33 elder AD subjects and 13 young cognitively normal controls. The $R^2$ for both calibrations is within tolerance (> 0.7). The ratio of standard deviations between methods in the PiB control group is 5.30/4.33 = 1.22. The ratio of standard deviations between methods in the FBP control group is 10.57/2.68 = 3.94.

Level 2: PiB Calibration using Pittsburgh Data

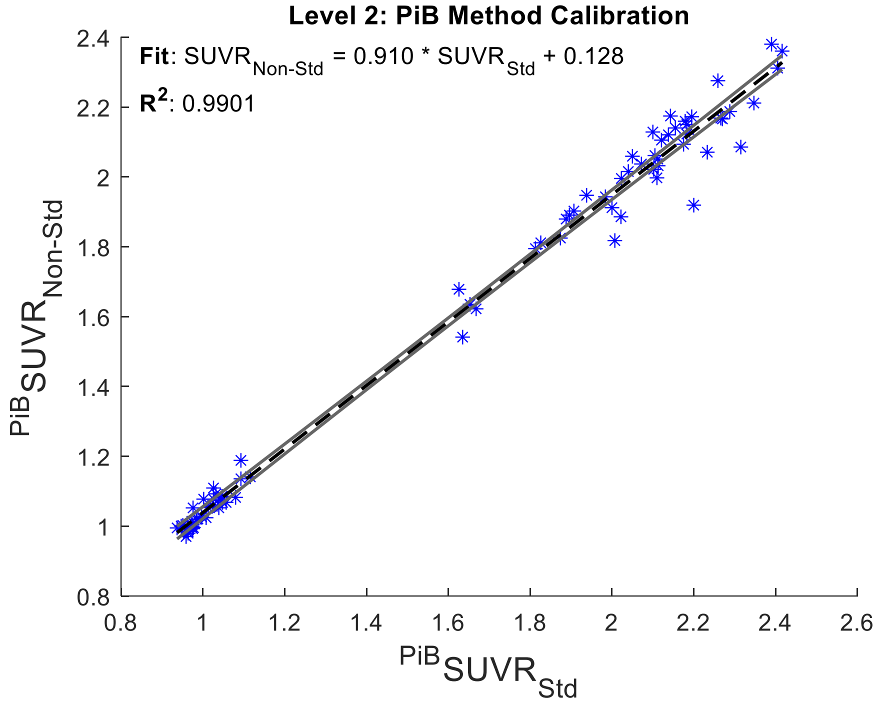


Figure S2 – Level 2 Centiloid calibration analysis for PiB using published Pittsburgh calibration data. The regression equation is substituted into the published Level 1 Centiloid equation to find:

$$CL = 100 * ({}^{PiB}SUVR_{Non-Std} - 1.047)/0.971$$

Level 2: FBP Calibration using Avid Data

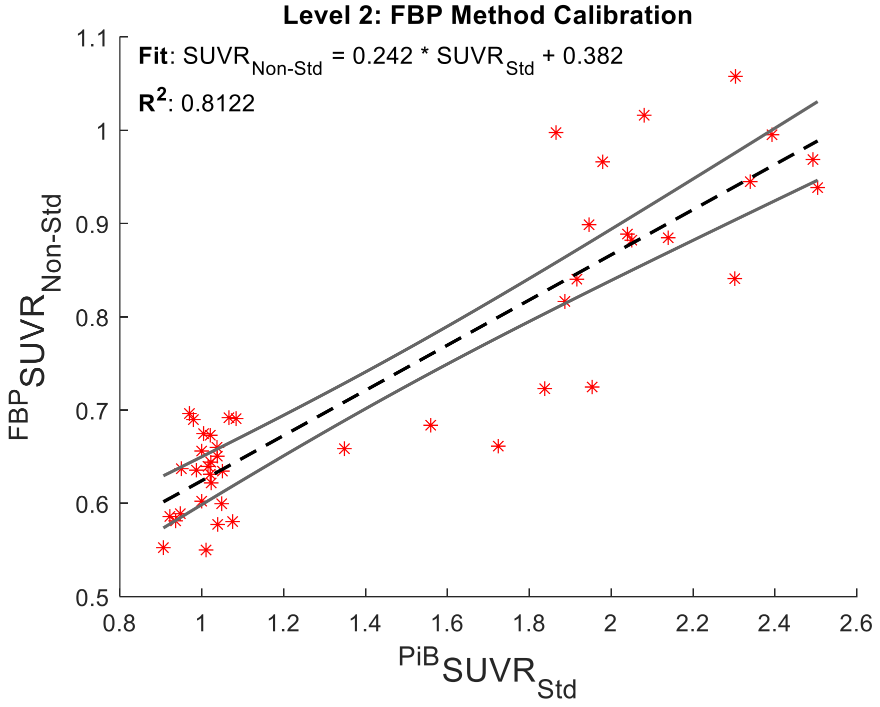


Figure S3 – Level 2 Centiloid calibration analysis for FBP using published Avid calibration data. The regression equation is substituted into the published Level 1 Centiloid equation to find:

$$CL = 100 * ({}^{FBP}SUVR_{Non-Std} - 0.626)/0.258$$

Matched Pair Demographics

| | PiB | FBP | P |
|---|---|---|---|
| Sample Size (n) | 66 | 66 | N/A |
| Age (yrs) | 44.8 (SD = 8.9) | 49.3 (SD = 6.9) | 0.002** |
| Female (%) | 35 (53%) | 22 (33.3%) | 0.022* |
| Aβ + (%) | 45 (68.2%) | 46 (69.7%) | 0.852 |
| APOE e4 Carriers (%) | 13 (19.7%) | 15 (22.7%) | 0.673 |

Table S3 – Demographics for CL-matched participants used in multivariate linear model.

SUVR and Gaussian Mixture Model Cutoffs

| **PiB** | **Mean SUVR (Aβ- group)** | **Mean SUVR (Aβ+ group)** | **GMM-derived SUVR Cutoff** |
|---|---|---|---|
| **Global Cortex** | 1.07 (SD=0.06) | 1.58 (SD=0.29) | 1.18 |
| **PET-based Striatum** | 1.09 (SD=0.18) | 1.98 (SD=0.37) | 1.12 |
| **MRI-based Striatum** | 1.04 (SD=0.14) | 1.64 (SD=0.27) | 1.11 |
| **Caudate** | 0.96 (SD=0.16) | 1.29 (SD=0.28) | 1.16 |
| **Putamen** | 1.13 (SD=0.12) | 1.80 (SD=0.32) | 1.22 |
| **Accumbens** | 1.00 (SD=0.15) | 1.69 (SD=0.30) | 1.08 |

| **FBP** | **Mean SUVR (Aβ- group)** | **Mean SUVR (Aβ+ group)** | **GMM-derived SUVR Cutoff** |
|---|---|---|---|
| **Global Cortex** | 0.63 (SD=0.03) | 0.83 (SD=0.10) | 0.75 |
| **PET-based Striatum** | 0.60 (SD=0.06) | 0.84 (SD=0.11) | 0.80 |
| **MRI-based Striatum** | 0.58 (SD=0.06) | 0.75 (SD=0.08) | 0.81 |
| **Caudate** | 0.55 (SD=0.08) | 0.64 (SD=0.09) | 0.79 |
| **Putamen** | 0.63 (SD=0.05) | 0.83 (SD=0.09) | 0.77 |
| **Accumbens** | 0.57 (SD=0.06) | 0.77 (SD=0.10) | 0.76 |

Table S4 – Average SUVR and Gaussian mixture model-derived cutoffs for PiB (top) and FBP (bottom).
